# Association of Visual Impairment with Brain Structure

**DOI:** 10.1101/2021.01.09.21249189

**Authors:** Zhuoting Zhu, Wenyi Hu, Huan Liao, Danli Shi, Zachary Tan, Yifan Chen, Xianwen Shang, Yu Huang, Xueli Zhang, Yu Jiang, Wei Wang, Xiaohong Yang, Mingguang He

## Abstract

**Objective:** To investigate the association of visual impairment (VI) with brain structures in the UK Biobank Study.

**Methods:** The UK Biobank Study is a large prospective study that recruited more than 500,000 participants aged 40-69 from 2006 to 2010 across the UK. Visual acuity (VA) of worse than 0.3 LogMAR units (Snellen 20/40) was defined as VI. Structural magnetic resonance imaging (MRI) data were obtained using a 3.0-T MRI imager. Volumetric measures of five global brain volumes (total brain volume, total grey matter, total white matter, cerebrospinal fluid (CSF), brain stem) and the volumes of seven specific brain region (thalamus, caudate nucleus, basal ganglia, pallidum, hippocampus, amygdala and nucleus accumbens) were included in the present analysis. Multivariable linear regression was used to investigate the association of VI with global and specific brain volumes.

**Results:** A total of 8976 participants free of neurological disorders at baseline assessment were included for the present analysis. The prevalence of VI was 0.02% (n=181). After adjusting for a range of cofounding factors, VI was significantly associated with decreased volumes of the total brain (β = -0.12, 95% confidence interval (CI) -0.23 to 0.00, P = 0.049), thalamus (β = -0.16, 95% CI -0.18 to -0.04, P = 0.010), caudate nucleus (β = -0.14, 95% CI -0.27 to 0.00, P = 0.046), pallidum (β = -0.15, 95% CI-0.27 to -0.02, P = 0.028) and amygdala (β = -0.18, 95% CI -0.31 to -0.04, P = 0.012).

**Interpretation:** We found that VI is associated with a decrease in total brain volumes and the volumes of specific brain regions implicated in neurodegenerative diseases.

## Introduction

Neurodegenerative diseases are a major cause of cognitive impairment and mortality in the ageing populations^1^. The two most common neurodegenerative diseases, Alzheimer’s disease (AD) and Parkinson’s disease (PD) affect nearly 50 million people worldwide^2, 3^. With the growth of the world population and the increase in life expectancy, the disease burden of neurodegenerative diseases is estimated to increase substantially in the near future.^3,4^ However, very few effective treatments for neurodegenerative diseases have been developed so far^5^, highlighting the importance of further investigating the underlying mechanisms of these diseases to facilitate the development of strategies for prevention and early detection.

As a projection of the central nerve system (CNS) via the optic nerve, the retina has been described as a “window to the brain”^6^. Additionally, growing evidence has reported strong associations of visual impairment (VI) with AD^7^ and PD^8^. Nevertheless, the mechanisms underlying the associations between VI and neurodegenerative diseases remains unknown. It has been widely recognized that structural changes in the brain could play important roles in the development and progression of neurodegenerative diseases^9-12^. Some studies have reported that structural changes in the brain, for example, decreased grey matter in the medial temporal lobe, may even precede the symptom onset of neurodegenerative diseases^13,14^. Therefore, a better understanding of the relationship between VI and brain structure may provide insights into the mechanisms behind the associations between VI and neurodegenerative diseases. To the best of our knowledge, no study to date has investigated the association between VI and brain structure in the middle-aged and elderly population.

Therefore, we aim to investigate the association of VI with brain structure in a large-scale community-based study.

## Methods

### Participants

The UK Biobank is a large prospective study with more than 500,000 enrolled in 2006-2010 and participants 40 to 69 years of age attended one of 22 assessment centers across the UK^15^. In late 2009, ocular assessments, including visual acuity (VA), refraction, intraocular pressure (IOP), corneal hysteresis (CH), optical coherence tomography (OCT) and fundus photography, were introduced to 6 centers across the UK, taken by approximately 117,175 participants^16^. In 2014, 100,000 original participants in the UK Biobank study were invited back for brain, heart and body imaging^17^. Participants (n=9310) with both VA testing and structural brain MRI available are included in the present study. For analysis of brain structure, a total of 217 individuals with self-reported neurological disorders (Supplement Table 1) and 117 with brain volumes of more than 4 standard deviations (SD) from mean values were excluded, resulting in a final sample of 8976 participants for analysis.

### Standard protocol approvals, registrations, and patient consents

Access to the UK Biobank data was granted after registration. The application ID was 62443. The UK Biobank has obtained Research Tissue Bank approval from its Research Ethics Committee recommended by the National Research Ethics Service (reference 11/NW/0382). Participants provided written informed consent.

### Visual Acuity Testing

Participants were tested for habitual VA based on a logarithm of the minimum angle of resolution (LogMAR) chart. The test was performed with participants at a distance of 4 meters with optical correction, if any, or at 1 meter if not being able to read. The right eye was measured first. Participants were asked to read the letter sequentially from the top to the smallest-sized letter they could reliably identify. The test was terminated if 2 letters were identified incorrectly and the total number of letters correctly identified was converted to LogMAR VA^16^. VA worse than 0.3 LogMAR units (Snellen 20/40) was defined as VI.

### Structural MRI

Structural MRI imaging data was obtained by a 3.0-T MRI imager (Siemens Skyra, Siemens Healthcare, Erlangen, Germany) with a standard 32-channel radiofrequency receiver head coil. T1-weighted structural imaging was performed in sagittal orientation by using a three-dimensional magnetization-prepared rapid acquisition with gradient echo sequence with a resolution of 1 × 1 × 1 mm, a field-of-view of 208 × 256 × 256 matrix and a duration of 5 minutes^17^. The processing of initially released data was performed by using FSL packages (the FMRIB Software Library, Oxford, England). Structural imaging segmentation into grey matter, white matter and cerebrospinal fluid (CSF) was applied by using FAST (Automated Segmentation Tool in FMRIB) and subcortical structures were modelled by using FIRST (FMRIB’s Integrated Registration and Segmentation Tool)^18^. MRI protocols, imaging processing and quality control are described in detail elsewhere^17, 18^. Data output by processing pipeline were used in the present study. A total of five global brain volumes (total brain volume, total grey matter, total white matter, CSF, brain stem) and volumes of seven specific brain regions including thalamus, caudate nucleus, basal ganglia, pallidum, hippocampus, amygdala and nucleus accumbens were included. Brain volumes were standardized to z-scores.

### Covariates

Factors known to be associated with brain structure were included as covariates in the present analysis^19-22^, including age at baseline assessment, sex (male/female), ethnicity (Caucasian and non-Caucasian), obtainable education (college or university degree, and others), smoking status (current/previous and never), diabetes mellitus (yes/no), hypertension (yes/no), major depression (yes/no) and body mass index (BMI).

Age at baseline assessment was categorized into three groups: 40 to 49, 50 to 59, and 60 to 69 years old. Diabetes was defined to include those participants who had self-reported or doctor-diagnosed diabetes mellitus, were taking anti-hyperglycemic medications or using insulin or had a glycosylated hemoglobin level of ≥6.5%. Hypertension was defined to include those participants who had self-reported, or doctor-diagnosed hypertension, were taking antihypertensive drugs or had a systolic blood pressure of at least 130 mmHg or a diastolic blood pressure of at least 80 mmHg averaged over two measurements. Self-reporting and/or the score on the Patient Health Questionnaire (PHQ, the first two items) of at least 3 were used to identify participants with depression^15^. BMI was calculated as the weight in kilograms divided by height in meters squared (kg/m^2^).

### Statistical analysis

Baseline characteristics stratified by VI were reported as number and proportion for categorical data, mean and standard deviation for continuous variables. Comparisons of categorical variables were examined by chi-square test and continuous variables by unpaired t-test. Multivariable linear regression was applied to model associations of VI with total and specific brain volumes. We firstly adjusted for age and gender (model I). The Benjamin-Hochberg procedure was employed to control the false discovery rate (FDR) at the level of 5%. Ethnicity, obtainable education, smoking status, diabetes mellitus, hypertension, major depression and BMI were further adjusted for if the model I achieved FDR < 5% significance. To reduce the prominent effect of ageing on brain structure, we performed sensitivity analysis controlling for age as a continuous variable and investigated the non-linear effects of age with adjustments of age and age squared. Analyses were performed using Stata version 13 (version 14.0; StataCorp).

### Data Availability Statement

All bona fide researchers could apply to UK Biobank resources for health-related research with benefit to public health. (see ukbiobank.ac.uk/register-apply).

## Results

### Study Sample

The present analysis included a sample of 8976 participants free of neurological disorders. The baseline characteristics of the participants are shown in Table 1. The prevalence of VI was 0.02% (n=181). Visually impaired participants were more likely to be older and of non-Caucasian ethnicity. There were no significant differences in the proportion of gender, obtainable education, smoking status, diabetes mellitus, hypertension, major depression and in the value of BMI between VI and non-VI groups.

**Table 1.**
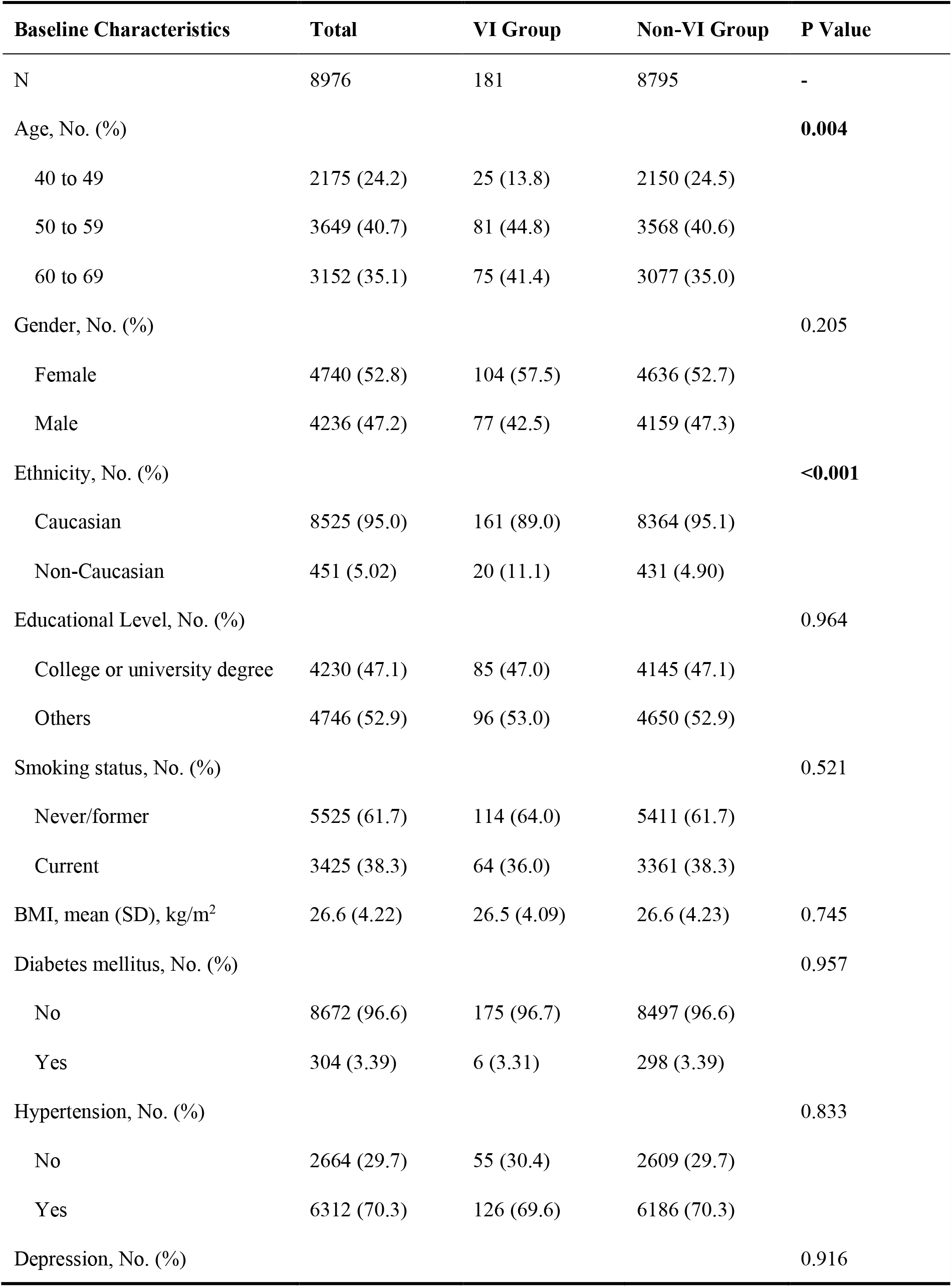
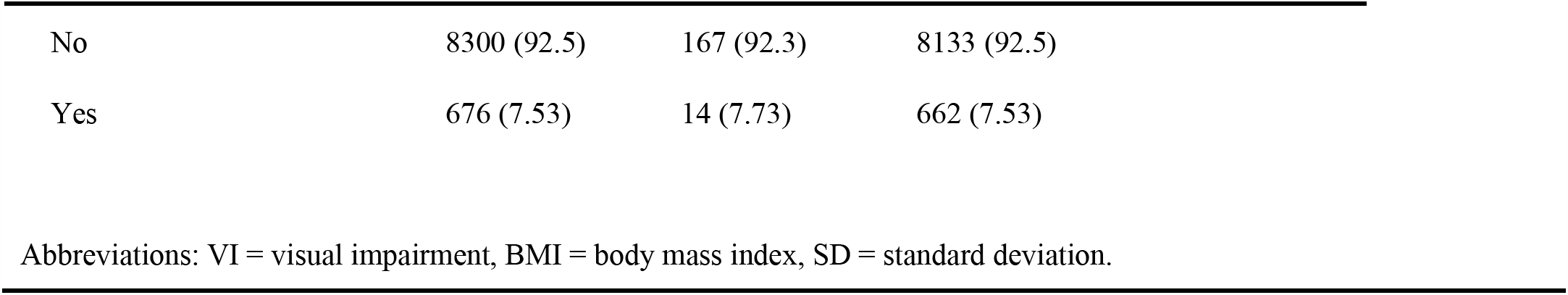
Baseline Characteristics of Study Participants Stratified by Visual Impairment Status.

| Baseline Characteristics | Total | VI Group | Non-VI Group | P Value |
| --- | --- | --- | --- | --- |
| N | 8976 | 181 | 8795 | - |
| Age, No. (%) |  |  |  | **0.004** |
| 40 to 49 | 2175 (24.2) | 25 (13.8) | 2150 (24.5) |  |
| 50 to 59 | 3649 (40.7) | 81 (44.8) | 3568 (40.6) |  |
| 60 to 69 | 3152 (35.1) | 75 (41.4) | 3077 (35.0) |  |
| Gender, No. (%) |  |  |  | 0.205 |
| Female | 4740 (52.8) | 104 (57.5) | 4636 (52.7) |  |
| Male | 4236 (47.2) | 77 (42.5) | 4159 (47.3) |  |
| Ethnicity, No. (%) |  |  |  | **<0.001** |
| Caucasian | 8525 (95.0) | 161 (89.0) | 8364 (95.1) |  |
| Non-Caucasian | 451 (5.02) | 20 (11.1) | 431 (4.90) |  |
| Educational Level, No. (%) |  |  |  | 0.964 |
| College or university degree | 4230 (47.1) | 85 (47.0) | 4145 (47.1) |  |
| Others | 4746 (52.9) | 96 (53.0) | 4650 (52.9) |  |
| Smoking status, No. (%) |  |  |  | 0.521 |
| Never/former | 5525 (61.7) | 114 (64.0) | 5411 (61.7) |  |
| Current | 3425 (38.3) | 64 (36.0) | 3361 (38.3) |  |
| BMI, mean (SD), kg/m<sup>2</sup> | 26.6 (4.22) | 26.5 (4.09) | 26.6 (4.23) | 0.745 |
| Diabetes mellitus, No. (%) |  |  |  | 0.957 |
| No | 8672 (96.6) | 175 (96.7) | 8497 (96.6) |  |
| Yes | 304 (3.39) | 6 (3.31) | 298 (3.39) |  |
| Hypertension, No. (%) |  |  |  | 0.833 |
| No | 2664 (29.7) | 55 (30.4) | 2609 (29.7) |  |
| Yes | 6312 (70.3) | 126 (69.6) | 6186 (70.3) |  |
| Depression, No. (%) |  |  |  | 0.916 |

|  |  |  |  |
| --- | --- | --- | --- |
| No | 8300 (92.5) | 167 (92.3) | 8133 (92.5) |
| Yes | 676 (7.53) | 14 (7.73) | 662 (7.53) |

Abbreviations: VI = visual impairment, BMI = body mass index, SD = standard deviation.

### Brain Structure

Table 2 shows global brain volumes and the volumes of specific brain regions stratified by VI status. Except for the cerebrospinal fluid, visually impaired participants had lower volumes of other global and all specific regions of the brain.

**Table 2.** Brain Volume of Study Participants Stratified by Visual Impairment Status.

| Brain Volume <sup>a</sup> | VI Group | Non-VI Group | P Value |
| --- | --- | --- | --- |
| Global volume |  |  |  |
| Total brain volume | **-0.27 (-0.41, -0.13)** | **0.01 (-0.02, 0.03)** | **<0.001** |
| Total grey matter | **-0.28 (-0.42, -0.13)** | **0.01 (-0.02, 0.03)** | **<0.001** |
| Total white matter | **-0.23 (-0.37, -0.09)** | **0.00 (-0.02, 0.03)** | **0.002** |
| Brain stem | **-0.20 (-0.34, -0.06)** | **0.00 (-0.02, 0.02)** | **0.008** |
| Cerebrospinal fluid | 0.08 (-0.07, 0.23) | -0.00 (-0.02, 0.02) | 0.300 |
| Specific region volume |  |  |  |
| Thalamus | **-0.31 (-0.45, -0.17)** | **0.01 (-0.01, 0.03)** | **<0.001** |
| Caudate nucleus | **-0.22 (-0.36, -0.08)** | **0.00 (-0.02, 0.03)** | **0.003** |
| Putamen nucleus | **-0.26 (-0.40, -0.11)** | **0.01 (-0.02, 0.03)** | **<0.001** |
| Pallidum | **-0.23 (-0.37, -0.09)** | **0.00 (-0.02, 0.03)** | **0.002** |
| Hippocampus | **-0.20 (-0.34, -0.05)** | **0.00 (-0.02, 0.02)** | **0.007** |
| Amygdala | **-0.23 (-0.37, -0.09)** | **0.00 (-0.02, 0.03)** | **0.002** |
| Nucleus accumbens | **-0.23 (-0.37, -0.09)** | **0.00 (-0.02, 0.03)** | **0.002** |

<sup>a</sup> Brain volumes were standardized to z-scores.

### Association of VI with Brain Structure

The associations of VI with multiple brain structures are reported in Table 3. The age- and gender-adjusted regression models demonstrated that VI was associated with lower volumes of the total brain (β = −0.17, 95% confidence interval (CI) −0.29 to-0.05, P = 0.020), total grey matter (β = −0.17, 95% CI −0.29 to −0.05, P = 0.021), total white matter (β = −0.16, 95% CI −0.27 to −0.04, P = 0.021), brain stem (β = −0.16, 95%CI −0.28 to −0.03, P = 0.021), thalamus (β = −0.19, 95% CI −0.31 to −0.07, P = 0.020), caudate nucleus (β = −0.17, 95% CI −0.30 to −0.03, P = 0.021), putamen nucleus (β =-0.16, 95% CI −0.28 to −0.03, P = 0.021), pallidum (β = −0.18, 95% CI −0.31 to −0.04, P = 0.021) and amygdala (β = −0.20, 95% CI −0.34 to −0.06, P = 0.020). In the full-adjusted models, VI had significant negative associations with the volumes of the total brain (β = −0.12, 95% confidence interval (CI) −0.23 to 0.00, P = 0.049), thalamus (β = −0.16, 95% CI −0.18 to −0.04, P = 0.010), caudate nucleus (β = −0.14,95% CI −0.27 to 0.00, P = 0.046), pallidum (β = −0.15, 95% CI −0.27 to −0.02, P =0.028) and amygdala (β = −0.18, 95% CI −0.31 to −0.04, P = 0.012).

**Table 3.** Associations of VI with Brain Volume.

| Brain Volume <sup>a</sup> | Age- and Gender-Adjusted Model |  | Multivariable Model <sup>c</sup> |  |
| --- | --- | --- | --- | --- |
| | $\beta$ (95% CI) | P Value <sup>b</sup> | $\beta$ (95% CI) | P Value |
| Global volume |  |  |  |  |
| Total brain | **-0.17 (-0.29, -0.05)** | **0.020** | **-0.12 (-0.24, -0.00)** | **0.049** |
| Total grey matter | **-0.16 (-0.29, -0.04)** | **0.021** | -0.11 (-0.23, 0.01) | 0.081 |
| Total white matter | **-0.15 (-0.27, -0.03)** | **0.021** | -0.12 (-0.24, 0.00) | 0.055 |
| Brain stem | **-0.16 (-0.29, -0.02)** | **0.021** | -0.12 (-0.25, 0.01) | 0.070 |
| Cerebrospinal fluid | 0.03 (-0.10, 0.15) | 0.794 | - | - |
| Specific region volume |  |  |  |  |
| Thalamus | **-0.20 (-0.33, -0.07)** | **0.020** | **-0.17 (-0.29, -0.04)** | **0.010** |
| Caudate nucleus | **-0.16 (-0.30, -0.03)** | **0.021** | **-0.14 (-0.28, -0.00)** | **0.046** |
| Putamen nucleus | **-0.16 (-0.29, -0.03)** | **0.021** | -0.12 (-0.25, 0.01) | 0.062 |
| Pallidum | **-0.17 (-0.31, -0.03)** | **0.021** | **-0.16 (-0.29, -0.02)** | **0.028** |
| Hippocampus | -0.13 (-0.26, 0.01) | 0.103 | - | - |
| Amygdala | **-0.20 (-0.34, -0.06)** | **0.020** | **-0.18 (-0.32, -0.04)** | **0.012** |
| Nucleus accumbens | -0.14 (-0.27, -0.00) | 0.086 | - | - |

<sup>a</sup> Brain volumes were standardized to z-scores.
<sup>b</sup> Corrected for multiple comparisons.
<sup>c</sup> Adjusted for age, sex, ethnicity, obtainable education, smoking status, diabetes mellitus, hypertension, major depression and body mass index.

### Sensitivity Analysis

Sensitivity analysis controlling for age as a continuous variable was performed. The significant associations of total brain volume or caudate volume with VI shown in the main analysis had only marginal significance. With respect to volumes of other global and specific regions, similar results to the main analysis were yielded (Supplement Table 2). After further adjustments for squared age, the results continued to remain similar (Supplement Table 2).

## Discussion

In the present analysis, we found that visually impaired individuals had lower volumes of total brain and multiple specific regions of brain structures, including the thalamus, caudate nucleus, pallidum and amygdala. Our findings provide insights into the mechanisms underlying the association between VI and neurodegenerative diseases.

To the best of our knowledge, this is the first study to date to report the association of VI with brain structure in the middle-aged and elderly participants. A previous study investigated the effects of congenital VI on brain structure in children^23^. They found defects in the development of the posterior tracts of the visual system and reduced volume of the thalamus in children with congenital VI. Limited numbers of studies have investigated the associations of ocular diseases with changes in brain structure^24-26^. Patients with high myopia showed altered grey matter volumes in terms of the visual pathway and limbic systems^24^. Wang et al.^25^ described altered brain structure (visual and non-visual cortex, lateral geniculate body and amygdala) in patients with glaucoma. Similarly, another study reported that the volume of the lateral geniculate body was negatively associated with the cup-disc ratio^26^. Consistent with previous findings, the present analysis observed atrophy of the visual pathway (thalamus) and limbic systems (amygdala) in the visually impaired participants. Intriguingly, atrophy of pallidum and caudate, which have well-established roles in motor regulation, was also observed in participants with VI. Study population (e.g., age range, ethnic background, sociodemographic characteristics), study design, and statistical analysis (e.g., statistical model, covariates adjusted in the model) might explain the discrepancy in results between our study and previous studies.

Although the mechanisms behind the association between VI and brain atrophy are not clear, several proposed mechanisms may provide explanations. It has been indicated that the pathogenesis of neurodegenerative diseases could affect both peripheral visual pathways and brain structure^27, 28^. The reduced brain volume in the hub of the visual pathway, the thalamus, may support this hypothesis^29, 30^. Besides, the reduction in visual inputs due to VI requires significant cognitive reserve to compensate for the impaired vision and hence may increase the cognitive burden, which could subsequently increase the vulnerability of the brain to neurodegenerative pathology. This hypothesis echoed the changes in brain structures implicated in the etiology of cognitive impairment and neurodegenerative diseases (total brain, amygdala, pallidum, caudate and thalamus) observed in visually impaired participants^28, 31-37^. Global brain atrophy has been widely observed in patients with neurodegenerative diseases^38^. The amygdala is important in processing and enhancing emotional memory and its atrophy explains the neuropsychological symptoms in AD patients^39^. Pallidum and caudate nucleus play important roles in the regulation of voluntary movements. Atrophies of these regions have been observed in neurodegenerative diseases characterized by predominantly motor symptoms, such as Parkinson’s disease and Huntington’s disease^40, 41^.

Alternatively, the association between VI and brain atrophy may be mediated by psychological dysregulations, such as depression and social isolation. Individuals with VI were found to be at higher risks of depression and social isolation^42, 43^. Indeed, brain volume changes in regions related to cognitive function and emotional control (e.g. hippocampus and amygdala) were previously reported in individuals with depression and social isolation^44, 45^. Although depression is the most important risk factor for dementia and Parkinson’s disease^46, 47^, the changes in brain volumes identified in the present study could not be entirely explained by depression, as the association between VI and brain volume reduction remained significant after adjusting for depression.

There are several strengths of our study, including the large-scale sample size, comprehensive MRI data on the total brain and specific brain structures, subjective measure of VA and comprehensive adjustments of confounding factors. However, our study also has some limitations. Firstly, due to the cross-sectional design of the study, we are unable to clarify the causality between VI and altered brain structures. Further longitudinal studies are warranted to investigate the association between VI and the progression of alterations in brain structure. Secondly, the participants recruited in the UK biobank do not represent the whole sampling population due to “healthy volunteer” selection bias^48^. Nevertheless, the investigation of the association between VI and brain structure does not require full representativeness and the results retain its robustness. Lastly, residual confounding could not be completely excluded.

## Conclusion

In conclusion, our study found that VI is associated with decreased volumes in the total brain and specific brain regions that are related to neurodegenerative diseases. Further studies are warranted to examine the longitudinal relationship between VI and brain structure alterations.

## Data Availability

All bona fide researchers could apply to UK Biobank resources for health-related research with benefit to public health. (see ukbiobank.ac.uk/register-apply). The application ID for the present study was 62443.

## Abbreviations and Acronyms

VI: visual impairment
VA: visual acuity
MRI: magnetic resonance imaging
CNS: central nervous system
AD: Alzheimer’s disease
PD: Parkinson’s disease
CSF: cerebrospinal fluid
BMI: body mass index
IOP: intraocular pressure
CH: corneal hysteresis
OCT: optical coherence tomography
SD: standard deviation
FDR: false discovery rate
PHQ: Patient Health Questionnaire
LogMAR: logarithm of the minimum angle of resolution
CI: confidence intervals

## Acknowledgements

The present work was supported by the Fundamental Research Funds of the State Key Laboratory of Ophthalmology, Project of Investigation on Health Status of Employees in Financial Industry in Guangzhou, China (Z012014075), Science and Technology Program of Guangzhou, China (202002020049). Prof. Mingguang He receives support from the University of Melbourne at Research Accelerator Program and the CERA Foundation. The Centre for Eye Research Australia receives Operational Infrastructure Support from the Victorian State Government. The sponsor or funding organization had no role in the design or conduct of this research.

## Author Contributions

Study concept and design: Zhu ZT, Hu WY, Liao H, He MG, Yang XH.

Acquisition, analysis, or interpretation: All authors.

Drafting of the manuscript: Zhu ZT, Hu WY.

Critical revision of the manuscript for important intellectual content: Tan Z, Chen YF, Shi DL, Wang W, He MG, Yang XH.

Statistical analysis: Zhu ZT, Shang XW.

Obtained funding: He MG, Yang XH.

Administrative, technical, or material support: Zhu ZT, Shang XW, Wang W, He MG, Yang XH.

Study supervision: Wang W, He MG, Yang XH.

