## Supplement Table 1 for "Association of Visual Impairment with Brain Structure"

Supplement Table 1. UK Biobank Code for Neurological Disorders Excluded from the Analysis

| **Neurological Disorders** | **UK Biobank Code** |
|---|---|
| Dementia or Alzheimer's disease | 1263 |
| Parkinson's disease | 1262 |
| Chronic degenerative neurological | 1258 |
| Guillain-Barré syndrome | 1256 |
| Multiple Sclerosis | 1261 |
| Other demyelinating disease | 1397 |
| Stroke or ischaemic stroke | 1081 |
| Brain cancer | 1032 |
| Brain haemorrhage | 1491 |
| Brain/intracranial abscess | 1245 |
| Cerebral aneurysm | 1425 |
| Cerebral palsy | 1433 |
| Encephalitis | 1246 |
| Epilepsy | 1264 |
| Head injury | 1266 |
| Infections of the nervous system | 1244 |
| Ischaemic stroke | 1583 |
| Meningeal cancer | 1031 |
| Meningioma (benign) | 1659 |
| Meningitis | 1247 |
| Motor Neuron Disease | 1259 |
| Neurological injury/trauma | 1240 |
| Spina bifida | 1524 |
| Subdural haematoma | 1083 |

| | |
|---|---|
| Subarachnoid haemorrhage | 1086 |
| Transient ischaemic attack | 1082 |
