## Supplement Table 2 for "Association of Visual Impairment with Brain Structure"

Supplement Table 2. Sensitivity Analysis for β Coefficients for the Association between VI and Brain Structure

| | Multivariable Model [a] | | Multivariable Model [b] | |
|---|---|---|---|---|
| **Brain Structure** | ***β* (95% CI)** | **P Value** | ***β* (95% CI)** | **P Value** |
| Global volume | | | | |
| Total brain | -0.10 (-0.22, 0.02) | 0.106 | -0.10 (-0.22, 0.02) | 0.092 |
| Total grey matter | -0.08 (-0.20, 0.04) | 0.196 | -0.08 (-0.20, 0.04) | 0.173 |
| Total white matter | -0.10 (-0.22, 0.02) | 0.088 | -0.11 (-0.23, 0.01) | 0.078 |
| Brain stem | -0.11 (-0.25, 0.02) | 0.092 | -0.12 (-0.25, 0.01) | 0.071 |
| Specific region volume | | | | |
| Thalamus | **-0.14 (-0.27, -0.02)** | **0.027** | **-0.15 (-0.27, -0.02)** | **0.022** |
| Caudate nucleus | -0.14 (-0.28, 0.00) | 0.057 | -0.13 (-0.27, 0.01) | 0.059 |
| Putamen nucleus | -0.10 (-0.22, 0.03) | 0.136 | -0.10 (-0.23, 0.02) | 0.112 |
| Pallidum | **-0.14 (-0.28, -0.00)** | **0.048** | **-0.15 (-0.28, -0.01)** | **0.039** |
| Amygdala | **-0.18 (-0.32, -0.04)** | **0.014** | **-0.18 (-0.32, -0.04)** | **0.013** |

[a] Adjusted for age as a continuous variable, sex, ethnicity, obtainable education, smoking status, diabetes mellitus, hypertension, major depression and body mass index.

[b] Adjusted for age as a continuous variable, squared age, sex, ethnicity, obtainable education, smoking status, diabetes mellitus, hypertension, major depression and body mass index.
